# Assessing the potential cost-effectiveness of centralized vs point-of-care testing for hepatitis C virus in Pakistan: a model-based comparison

**DOI:** 10.1101/2022.03.31.22273228

**Authors:** Joseph B. Babigumira, James K. Karichu, Samantha Clark, Mindy M. Cheng, Louis P. Garrison, Maciej B. Maniecki, Saeed S. Hamid

## Abstract

**Background:** Pakistan has a hepatitis C virus (HCV) infection prevalence of 6–9% and aims to achieve World Health Organization (WHO) targets for elimination of HCV by the year 2030 through scaling HCV diagnosis and accelerating access to care. The clinical and economic benefits of various HCV testing strategies have not yet been evaluated in Pakistan.

**Objective:** To evaluate the potential cost-effectiveness of a reference laboratory-based (CEN) confirmatory testing approach vs a molecular near-patient point-of-care (POC) confirmatory approach to screen the general population for HCV in Pakistan.

**Methods:** We developed a decision-analytic model comparing HCV testing under two scenarios: screening with an anti-HCV antibody test (Anti-HCV) followed by either POC nucleic acid testing (NAT) (Anti-HCV-POC), or reference laboratory NAT (Anti-HCV-CEN), using data from published literature, the Pakistan Ministry of Health, and expert judgment. Outcome measures included: number of HCV infections identified per year, percentage of individuals correctly classified, total costs, average costs per individual tested, and cost-effectiveness. Sensitivity analysis was also performed.

**Results:** At a national level for a tested population of 25 million, the Anti-HCV-CEN strategy would identify 142,406 more HCV infections in one year and increase correct classification of individuals by 0.57% compared with the Anti-HCV-POC strategy. The total annual cost of HCV testing was reduced using the Anti-HCV-CEN strategy by $7.68 million ($0.31 per person). Thus, incrementally, the Anti-HCV-CEN strategy costs less and identifies more HCV infections than Anti-HCV-POC.

**Conclusions:** Anti-HCV-CEN would provide the best value for money when scaling up HCV testing in Pakistan.

**Significance statement:**

- Hepatitis C virus (HCV) infection constitutes a major medical and public health burden in Pakistan
- Widespread testing is important to identify those that are chronically infected in order to link them to treatment services
- The optimal and most cost-effective testing approach to scale up HCV testing to support elimination efforts in Pakistan has not been established
- High throughput reference laboratory testing would provide the best value for money when scaling-up HCV testing in Pakistan

## Introduction

Hepatitis C virus (HCV) infection constitutes a major medical and public health burden worldwide, with an estimated 1% of the world population chronically infected.^1^ Over 80% of those affected live in low- and middle-income countries (LMICs).^2^ One-third of people with chronic, untreated HCV infection develop liver cirrhosis, and have higher morbidity and mortality rates.^3^

The advent of highly effective direct-acting antiviral agents (DAAs) has transformed the clinical care of HCV.^4-6^ DAAs are oral medications that can be instrumental in large-scale elimination efforts.^7^ Specifically, the World Health Organization (WHO) goals for HCV elimination include 95% of donation screening in a quality-assured manner by 2020, and 100% by 2030.^8^ However, limited access to diagnostic testing remains a major barrier.^8^ The prevalence of HCV infection in Pakistan is estimated to be 6–9% of the population,^9-12^ despite the availability of DAAs,^11^ in part because of the absence of a comprehensive, population-wide screening program. Several studies have estimated targets for screening, diagnosis, and treatment in Pakistan,^13-15^ and the Pakistani government has developed a policy framework based on the WHO guidelines to support rapid scale up of HCV testing.^16^

The current testing approach for HCV in Pakistan is achieved via a rapid anti-HCV antibody test (Anti-HCV) for screening, followed by reflex testing with a confirmatory nucleic acid test (NAT) if results indicate positivity.^17,18^ Rapid screening tests are typically home based, while NAT testing requires a clinic visit for venous blood collection. NAT testing can either be performed at a district laboratory or at a high-throughput reference laboratory, of which there is limited availability. As over 60% of the population lives in rural areas, large-scale HCV screening may be logistically challenging under this current approach.^19^

Decentralized molecular point-of-care (POC) testing (e.g. via the GeneXpert System^®^ in district hospitals) moves the site of testing closer to patients and is an alternative to reference laboratory-based NAT testing, offering reduced sample transportation time and faster results.^17,18^ However, POC NAT testing may be expensive to implement and poses logistical challenges, including a need for continuous electrical supply and adequate storage space for cartridges.^20,21^ Dried blood spot (DBS) testing from finger-prick samples solves many of these problems and facilitates access to testing as it is a low-resource option that requires no training for sample collection.^22^ Despite conditional recommendations from the WHO to use DBS specimens as an alternative to HCV NAT in settings where resources or expertise are limited,^3^ DBS testing has some drawbacks. Some studies have indicated that DBS tests have a higher limit of detection than serum and that variable DBS sample stability can impact quantification accuracy.^23-25^

The cobas^®^ Plasma Separation Card (PSC) offers another option; whilst currently approved only for the purpose of human immunodeficiency virus (HIV) viral load monitoring,^26^ the utility of the PSC for HCV testing has been recently demonstrated.^27,28^ The PSC has three sample collection ‘spots’ that allow three different tests to be performed with one card. Furthermore, the PSC separates serum from blood cells and can be collected at patients’ homes immediately after a positive anti-HCV test.

Samples remain stable for testing for up to 28 days at 18–45°C and up to 85% humidity,^29^ offering the potential ability to significantly expand access to HCV testing in remote areas while ensuring sample integrity. While both DBS and the PSC can facilitate decentralization of sample collection, the PSC retains the sample collection and transport advantages of DBS while maintaining sample stability and viability.^21,30^

A previous study has suggested that POC is a cost-effective option in Pakistan,^13^ but there is still a lack of health economic evidence on the optimal and most cost-effective approach to scale up HCV testing to support elimination efforts. Therefore, the primary purpose of this study was to evaluate the potential cost-effectiveness of high-throughput, reference laboratory-based confirmatory testing compared with a near-patient molecular POC approach to inform HCV testing scale-up plans in Pakistan.

### Methods

#### Analytic overview

A decision-analytic model (decision tree) was developed to compare chronic HCV testing under two scenarios: (1) screening with anti-HCV followed by POC NAT (Anti-HCV-POC) and (2) screening with anti-HCV followed by high-throughput, centralized reference laboratory-based NAT (Anti-HCV-CEN) (Figure 1). We used a governmental (formal healthcare sector) perspective, excluding third-party payer and patient out-of-pocket costs. We assessed costs and outcomes over a short-term time period, defined as the time from anti-HCV screening to HCV infection confirmation for anti-HCV-positive individuals.

**Figure 1.**
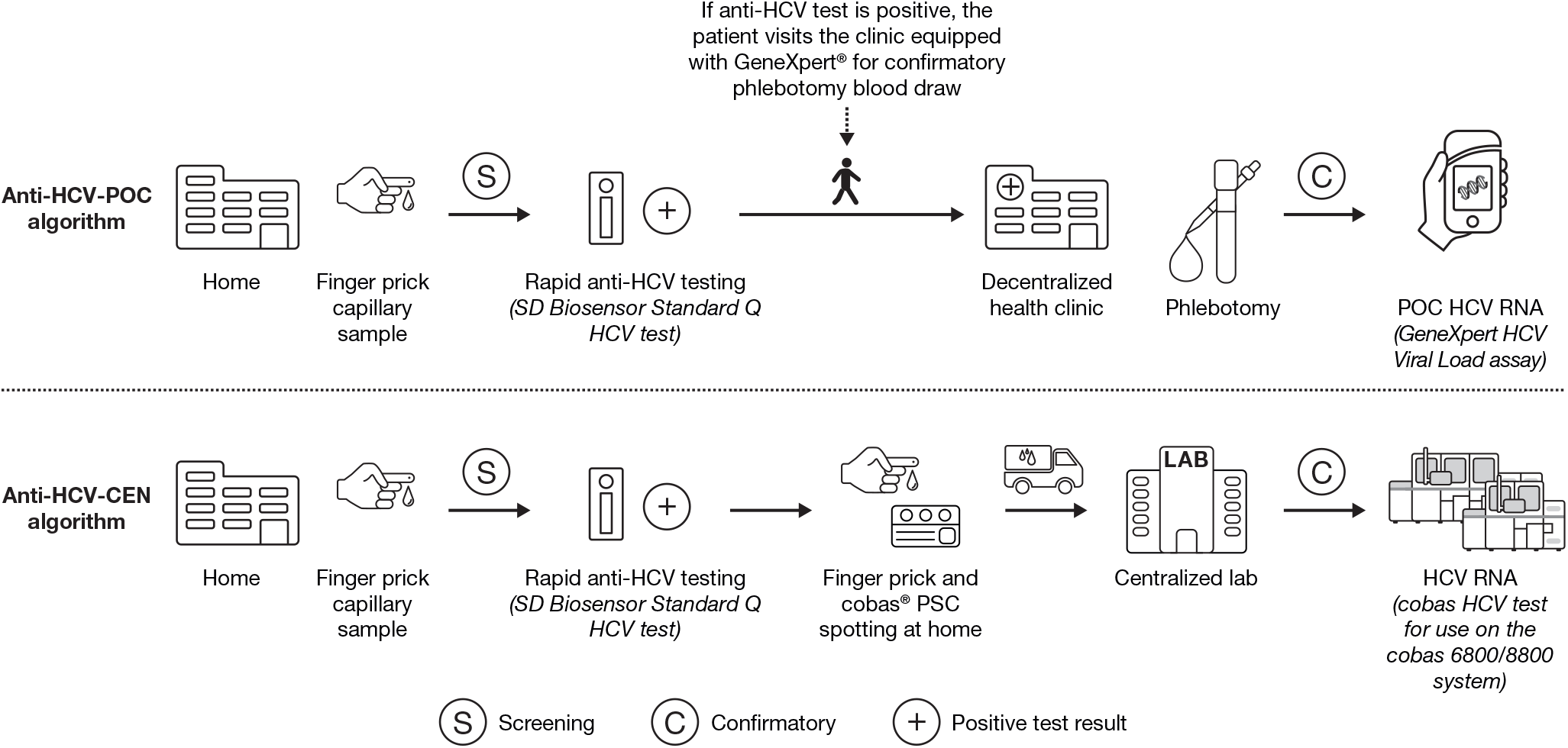
Anti-HCV-POC and Anti-HCV-CEN testing algorithms. CEN, central laboratory testing; HCV, hepatitis C virus; POC, point of care

#### Study population

The study population was the general testing population for chronic HCV in Pakistan, derived from published literature. Additional data related to costs and resource utilization were obtained from the Pakistan Ministry of Health (MOH), and expert judgement. As this research did not directly involve human subjects, informed consent was not required. The size of this population has been projected to be 25 million people annually, starting in 2018, to achieve chronic HCV elimination in Pakistan by 2030.^13^ Individuals were assumed to be initially screened at home, followed by POC NAT at nearby health facilities or followed by NAT at central laboratories.

#### Decision tree

The decision tree (Figure 2) modeled individuals in the testing population under the two scenarios (Anti-HCV-POC and Anti-HCV-CEN) by initially dividing them into HCV positive and HCV negative based on the population prevalence of chronic HCV in Pakistan. Depending on the test performance of the anti-HCV and NAT tests, individuals with HCV infection may test either positive (true positive [TP]) or negative (false negative [FN]), and individuals without HCV infection may test either positive (false positive [FP]) or negative (true negative [TN]). Individuals who tested TP or FP for HCV antibodies under the Anti-HCV-POC scenario were modeled to attend a nearby clinic for phlebotomy and POC NAT or were otherwise lost to follow-up (LTFU), implying non-compliance with a recommendation to receive confirmatory NAT. The model assumed no potential for LTFU under the Anti-HCV-CEN because PSC samples would be collected immediately after individuals receive a positive anti-HCV test at home and would be automatically transported to a reference laboratory for confirmatory NAT. Individuals who received NAT were assumed to generate either indeterminate tests or valid test results. Individuals who tested TP or TN were considered to have been correctly classified and thus exited the model. Individuals who tested either FN or FP, were LTFU, or had indeterminate test results were considered to have been incorrectly classified and thus exited the model.

**Figure 2.**
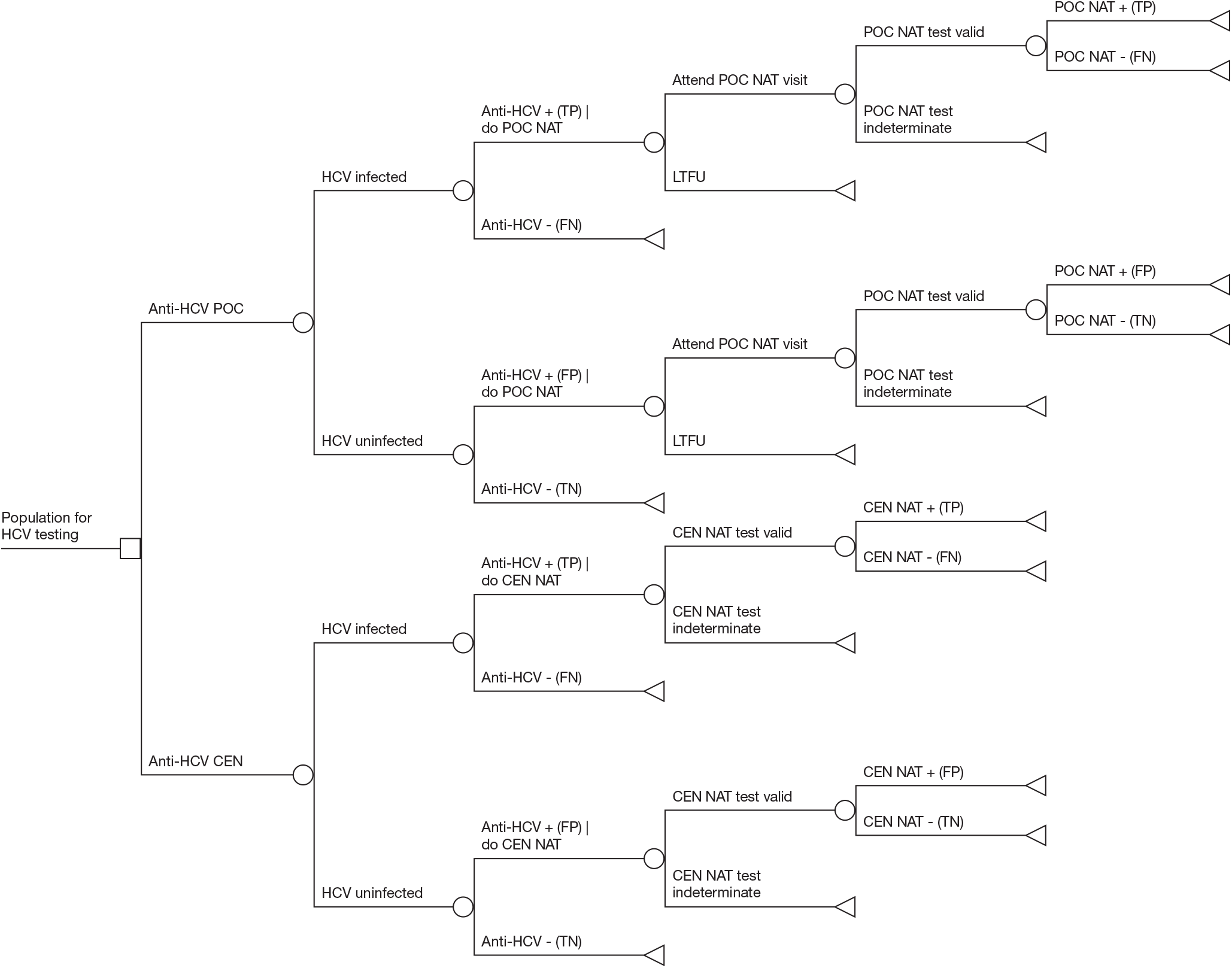
Decision-analytic model (decision tree) comparing Anti-HCV-CEN to Anti-HCV-POC in Pakistan. CEN, central laboratory testing; FP, false positive; FN, false negative; HCV, hepatitis C virus; LTFU, lost to follow-up; NAT, nucleic acid testing; POC, point of care; TP, true positive; TN, true negative

#### Probabilities

Table 1 includes a summary of the probabilities used in the model. The population prevalence of HCV in Pakistan was obtained from the published literature.^12,31^ The performance of the anti-HCV screening test was estimated based on a published review of multiple screening technologies, of which we used data on the performance of the SD Biosensor Standard Q HCV test on account of its superior performance.^32^ The performance of POC NAT testing was estimated based on multiple studies that reported the performance of the GeneXpert POC system under field conditions.^13,33^ The performance of central NAT testing was based on a synthesis of results of evaluations of the performance of the cobas^®^ 6800/8800 system under field conditions.^34^ The probability of being LTFU after screening positive for anti-HCV (i.e., not attending POC NAT testing) was estimated based on review of the HCV testing and treatment cascade,^35^ which found a wide variation in LTFU, including data from a study from Pakistan that reported that only 18% of those individuals who screened positive attended confirmatory testing.^36^ We used an estimate reported by the review that was based on multiple studies performed in the community and designed to improve the HCV care cascade,^35^ which will naturally follow efforts to increase testing for chronic HCV. The probabilities of obtaining indeterminate NAT results were obtained from field studies of the GeneXpert system^18^ and the cobas system.^37^

**Table 1.**
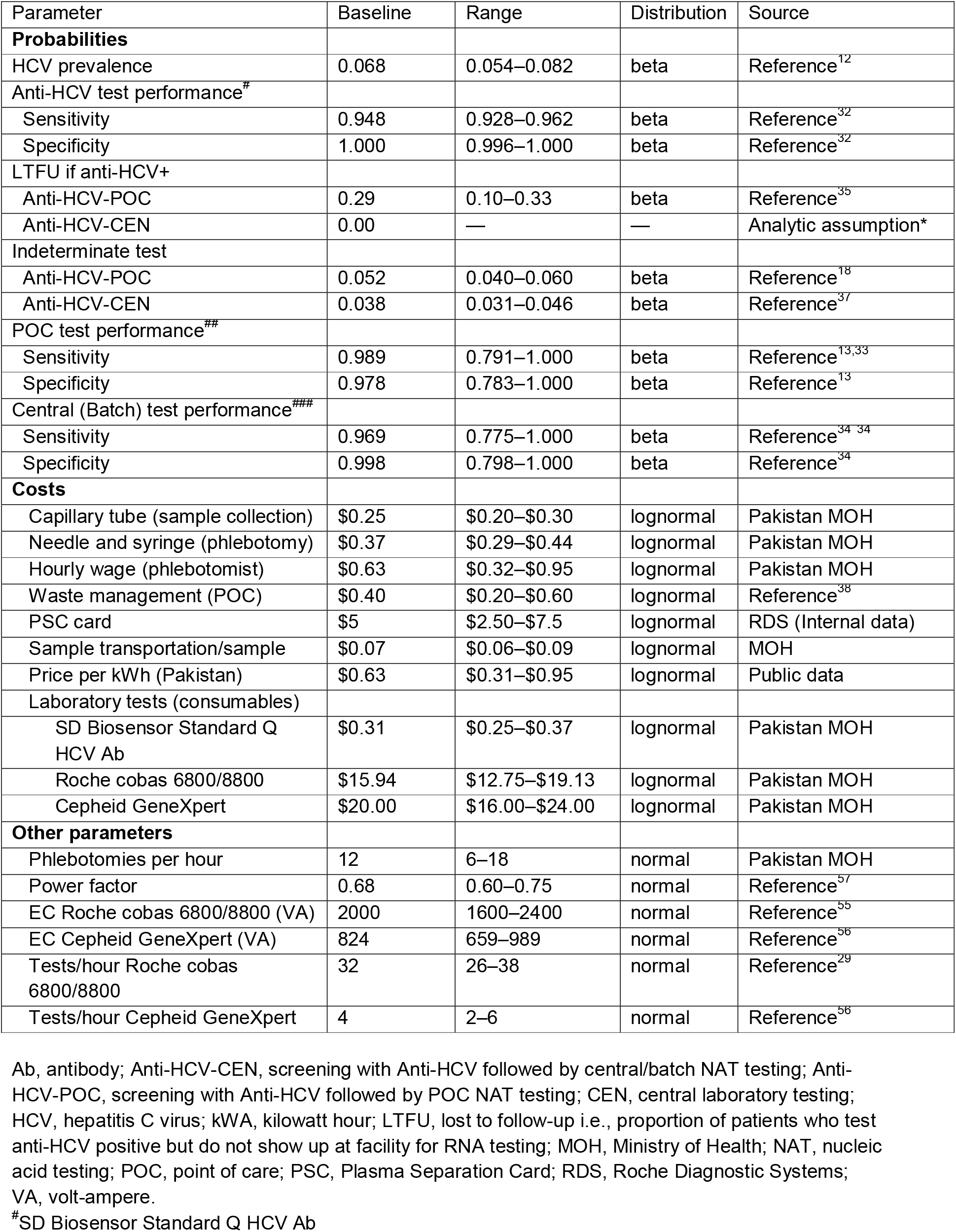
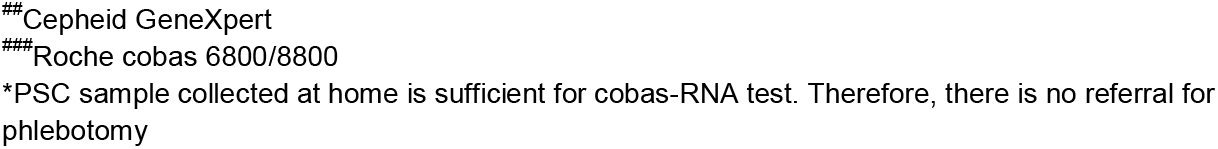
Parameters of the decision analytic model comparing HCV testing with Anti-HCV-CEN and Anti-HCV-POC in Pakistan

| Parameter | Baseline | Range | Distribution | Source |
| --- | --- | --- | --- | --- |
| **Probabilities** |  |  |  |  |
| HCV prevalence | 0.068 | 0.054–0.082 | beta | Reference<sup>12</sup> |
| Anti-HCV test performance<sup>#</sup> |  |  |  |  |
| Sensitivity | 0.948 | 0.928–0.962 | beta | Reference<sup>32</sup> |
| Specificity | 1.000 | 0.996–1.000 | beta | Reference<sup>32</sup> |
| LTFU if anti-HCV+ |  |  |  |  |
| Anti-HCV-POC | 0.29 | 0.10–0.33 | beta | Reference<sup>35</sup> |
| Anti-HCV-CEN | 0.00 | — | — | Analytic assumption* |
| Indeterminate test |  |  |  |  |
| Anti-HCV-POC | 0.052 | 0.040–0.060 | beta | Reference<sup>18</sup> |
| Anti-HCV-CEN | 0.038 | 0.031–0.046 | beta | Reference<sup>37</sup> |
| POC test performance<sup>##</sup> |  |  |  |  |
| Sensitivity | 0.989 | 0.791–1.000 | beta | Reference<sup>13,33</sup> |
| Specificity | 0.978 | 0.783–1.000 | beta | Reference<sup>13</sup> |
| Central (Batch) test performance<sup>###</sup> |  |  |  |  |
| Sensitivity | 0.969 | 0.775–1.000 | beta | Reference<sup>34, 34</sup> |
| Specificity | 0.998 | 0.798–1.000 | beta | Reference<sup>34</sup> |
| **Costs** |  |  |  |  |
| Capillary tube (sample collection) | \$0.25 | \$0.20–\$0.30 | lognormal | Pakistan MOH |
| Needle and syringe (phlebotomy) | \$0.37 | \$0.29–\$0.44 | lognormal | Pakistan MOH |
| Hourly wage (phlebotomist) | \$0.63 | \$0.32–\$0.95 | lognormal | Pakistan MOH |
| Waste management (POC) | \$0.40 | \$0.20–\$0.60 | lognormal | Reference<sup>38</sup> |
| PSC card | \$5 | \$2.50–\$7.5 | lognormal | RDS (Internal data) |
| Sample transportation/sample | \$0.07 | \$0.06–\$0.09 | lognormal | MOH |
| Price per kWh (Pakistan) | \$0.63 | \$0.31–\$0.95 | lognormal | Public data |
| Laboratory tests (consumables) |  |  |  |  |
| SD Biosensor Standard Q HCV Ab | \$0.31 | \$0.25–\$0.37 | lognormal | Pakistan MOH |
| Roche cobas 6800/8800 | \$15.94 | \$12.75–\$19.13 | lognormal | Pakistan MOH |
| Cepheid GeneXpert | \$20.00 | \$16.00–\$24.00 | lognormal | Pakistan MOH |
| **Other parameters** |  |  |  |  |
| Phlebotomies per hour | 12 | 6–18 | normal | Pakistan MOH |
| Power factor | 0.68 | 0.60–0.75 | normal | Reference<sup>57</sup> |
| EC Roche cobas 6800/8800 (VA) | 2000 | 1600–2400 | normal | Reference<sup>55</sup> |
| EC Cepheid GeneXpert (VA) | 824 | 659–989 | normal | Reference<sup>56</sup> |
| Tests/hour Roche cobas 6800/8800 | 32 | 26–38 | normal | Reference<sup>29</sup> |
| Tests/hour Cepheid GeneXpert | 4 | 2–6 | normal | Reference<sup>56</sup> |

Ab, antibody; Anti-HCV-CEN, screening with Anti-HCV followed by central/batch NAT testing; Anti-HCV-POC, screening with Anti-HCV followed by POC NAT testing; CEN, central laboratory testing; HCV, hepatitis C virus; kWh, kilowatt hour; LTFU, lost to follow-up i.e., proportion of patients who test anti-HCV positive but do not show up at facility for RNA testing; MOH, Ministry of Health; NAT, nucleic acid testing; POC, point of care; PSC, Plasma Separation Card; RDS, Roche Diagnostic Systems; VA, volt-ampere.
<sup>#</sup>SD Biosensor Standard Q HCV Ab

#### Costs

Costs were divided into five categories: (1) sample collection (phlebotomy for POC NAT and PSC card for central NAT); (2) waste management (incineration and waste transport) for POC; (3) sample transportation for central NAT; (4) consumption of electricity for NAT; and (5) testing consumables for all tests. The cost estimation did not include the fixed capital equipment or the maintenance costs of POC NAT or central NAT technology. Testing systems were assumed to be procured based on a reagent-rental model (i.e., the instrument is ‘free’ with the purchase of reagents) instead of direct purchase. The cost of phlebotomy was obtained from the Pakistan MOH and included estimates for the cost of blood collection tubes and syringes, as well as phlebotomists’ time, which was estimated from local hourly wages and estimated number of hourly phlebotomies performed. The per-test cost of waste management was obtained from the literature.^38^ The per-test cost of electricity was obtained from estimates of electricity consumption of the respective NAT technologies, number of tests per hour, and the per unit cost of electricity in Pakistan. The costs of testing consumables for all tests were based on the respective technologies and obtained from the Pakistan MOH. Costs obtained in local currency units (Pakistan Rupee) were converted into US Dollars using the official exchange rate of the State Bank of Pakistan (February 2021). Costs were not discounted given the short time period from an initial screening test to confirmatory test. Table 1 includes a summary of the parameters used for cost estimation.

#### Outcomes and cost-effectiveness

A baseline analysis was performed to estimate the following metrics under each testing scenario: (1) number of HCV infections identified per year; (2) percentage of individuals correctly classified (as either TP or TN); (3) total costs; (4) average costs per individual tested; and (5) cost-effectiveness in terms of cost per additional HCV infection identified.

#### Sensitivity analysis

A univariate sensitivity analysis was performed, re-estimating results with each parameter of the model at low and high values while holding all other parameters constant. The low and high values were 95% confidence intervals (CIs) when available and +/-20% for probabilities or +/-50% for costs when 95% CIs were unavailable. Monte Carlo simulation (1000 runs) was used to conduct probabilistic sensitivity analyses to assess overall parameter uncertainty in the model and further test the robustness of results. Baseline values were used as means, and standard errors were estimated assuming ranges were equivalent to 95% CIs (four times the standard error). Beta distributions were assumed for probabilities, lognormal distributions for costs, and normal distributions for counts. The analysis was performed using TreeAge Pro 2021 (TreeAge Software, LLC) and this report conforms to the Consolidated Health Economic Evaluation Reporting Standards (CHEERS) statement.^39^

## Results

### Baseline analysis

The results of the baseline analysis are shown in Table 2. Given an annual testing population of 25 million individuals, the Anti-HCV-CEN strategy identified 142,406 more HCV infections in 1 year compared with the Anti-HCV-POC strategy. The Anti-HCV-CEN strategy increased correct classification of individuals (TP and TN) by 0.57% compared with the Anti-HCV-POC strategy. The total annual cost of HCV testing in Pakistan was estimated to be $41.65 million ($1.67 per person) under Anti-HCV-CEN and $49.31 million ($1.97 per person) under Anti-HCV-POC. Anti-HCV-CEN reduced HCV testing costs in Pakistan by $7.68 million ($0.31 per person). In the incremental analysis, Anti-HCV-CEN was superior to Anti-HCV-POC (i.e., is less costly and identifies more HCV infections).

**Table 2.** Overall comparison of HCV testing with Anti-HCV-CEN and HCV testing with Anti-HCV-POC in Pakistan

|  | **Anti-HCV-POC** | **Anti-HCV-CEN** | **Incremental** |
| --- | --- | --- | --- |
| HCV infections identified per year | 1,359,892 | 1,502,298 | 142,406 |
| Individuals correctly classified | 98.64% | 99.21% | 0.57% |
| Total costs | \$49,331,577 | \$41,652,867 | -\$7,678,711 |
| Costs per individual screened | \$1.97 | \$1.67 | -\$0.31 |
| ICER (\$/additional HCV infection identified) | — | — | Anti-HCV-CEN is dominant* |

Anti-HCV-CEN, screening with anti-HCV followed by central/batch NAT testing; Anti-HCV-POC, screening with anti-HCV followed by POC NAT testing; CEN, central laboratory testing; HCV, hepatitis C virus; ICER, incremental cost-effectiveness ratio; NAT, nucleic acid testing; POC, point of care
\*Reduces costs and increases effectiveness, measured as HCV infections identified

### Sensitivity analysis

The incremental difference in HCV infections identified was most sensitive to the probability of LTFU (for POC confirmatory NAT). At 0% LTFU (i.e., all individuals attending a POC visit), Anti-HCV-CEN still identifies more infections, but the incremental difference in HCV infections identified decreases to 8693 (from 142,406 assuming 30% LTFU at baseline). At 33% LTFU, the incremental difference in HCV infections identified increases to 489,934 more HCV infections identified per year. The incremental costs were also most sensitive to the probability of LTFU (for POC confirmatory NAT), changing from -$0.49 (-$12,298,886 per year nationally) at 0% LTFU to $0.12 ($2,947,693 per year nationally) with a 33% rate of LTFU (Figure 3).

**Figure 3.**
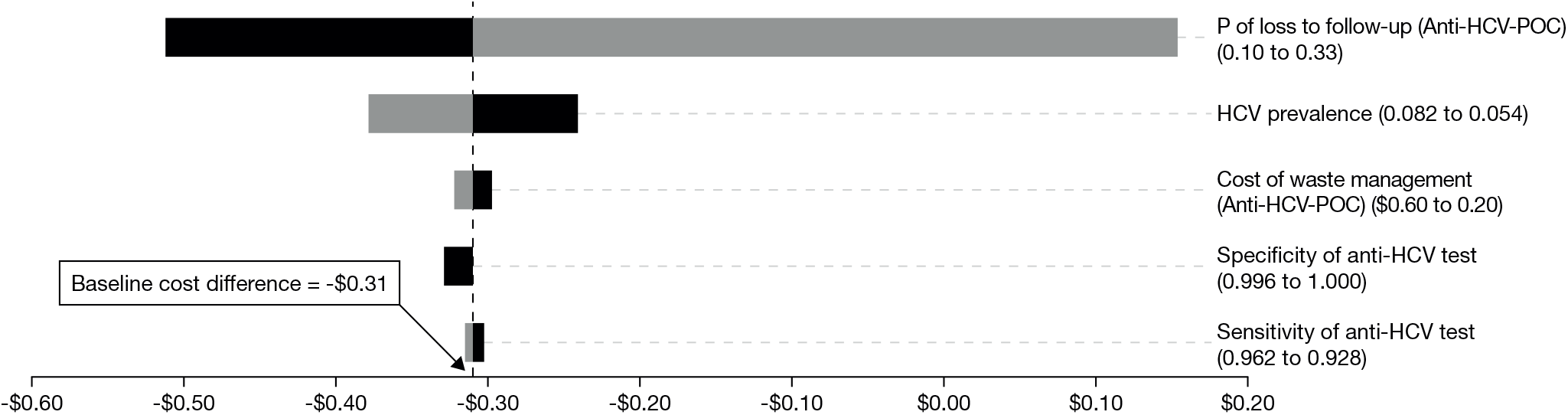
Tornado diagram of incremental costs comparing Anti-HCV-CEN to Anti-HCV-POC. The black bars represent minimum values while the grey bars represent high values (values are stated in parentheses) CEN, central laboratory testing; HCV, hepatitis C virus; P, probability; POC, point of care

The impact of parameter uncertainty in the model was represented graphically using PSA results in an incremental cost-effectiveness scatterplot (Figure 4). Anti-HCV-CEN is superior to Anti-HCV-POC (lower cost and greater number of HCV cases identified) in 71.1% of the simulations and inferior to Anti-HCV-POC (higher cost and fewer HCV cases identified) in 0.4% of the 1000 iterations of the Monte Carlo simulation.

**Figure 4.**
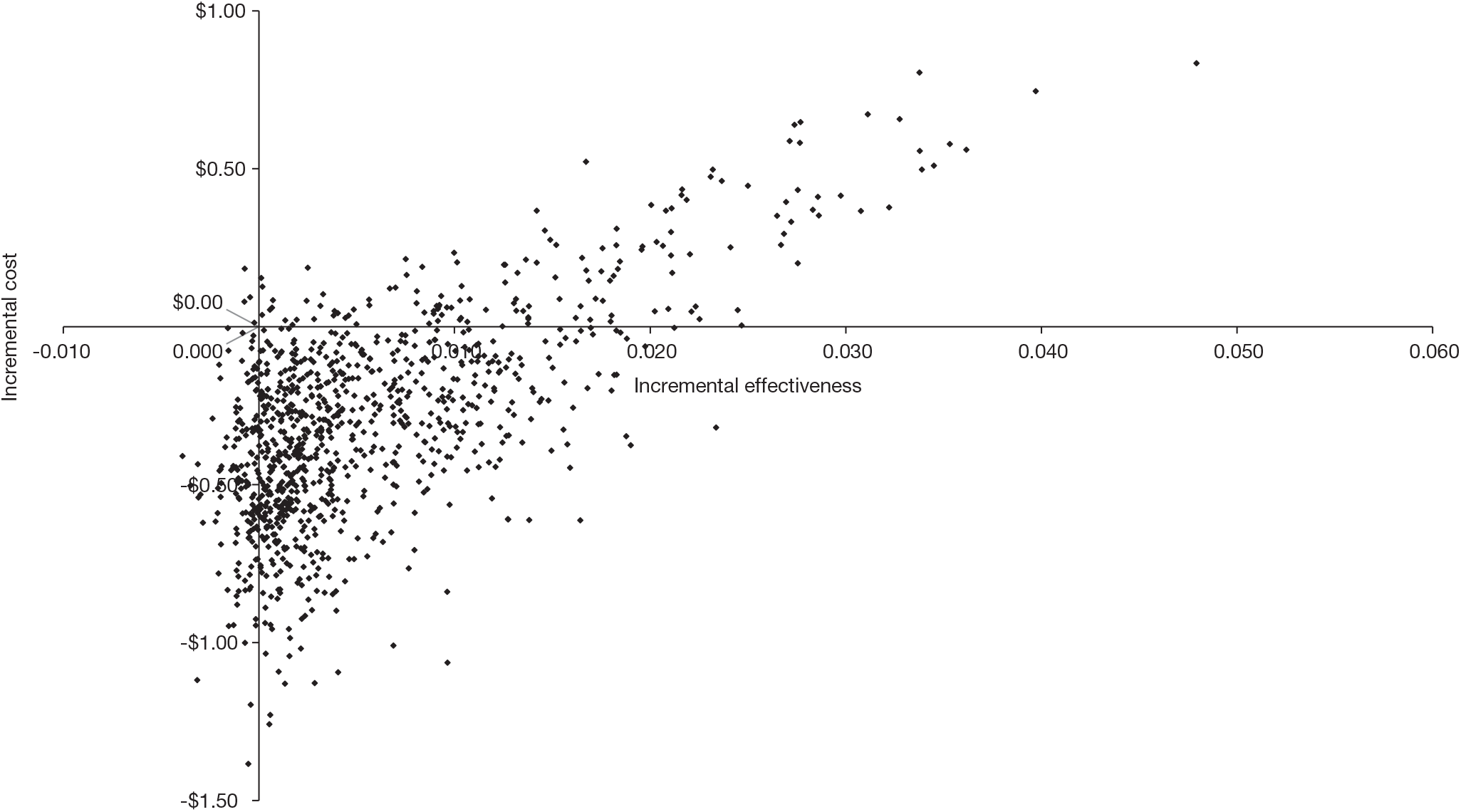
Incremental cost-effectiveness scatterplot comparing Anti-HCV-CEN to Anti-HCV-POC. CEN, central laboratory testing; HCV, hepatitis C virus; POC, point of care

## Discussion

We developed a decision-analytic model to compare chronic HCV testing under two scenarios: Anti-HCV-POC and Anti-HCV-CEN. The analysis projects that the Anti-HCV-CEN testing approach would identify more HCV infections and would increase the correct classification of individuals (TP and TN) at a lower cost compared with the Anti-HCV-POC strategy. Correctly identifying more chronically infected patients and providing the necessary treatment reduces onward disease transmission and contributes to Pakistan’s goal of eliminating HCV by the year 2030. Compared with the general population, patients with chronic HCV are at a significantly higher risk of developing costly HCV-related complications, including hepatic fibrosis, cirrhosis, and hepatocellular carcinoma.^40^

While the Anti-HCV-POC approach shortens the time between screening and treatment initiation,^36^ LTFU is common. The cobas PSC addresses challenges of LTFU whereby patients fail to adhere to recommended confirmatory follow-up testing. LTFU due to non-compliance with testing and treatment protocols has been described as a substantial challenge in Pakistan.^41^ To our knowledge, this is the first study in Pakistan to estimate the potential cost and accuracy of using the cobas PSC to collect samples for HCV testing. Given that over 60% of the Pakistan population resides in rural areas where access to testing is constrained,^19^ the cobas PSC has the potential to reduce LTFU and increase access to advanced molecular diagnostic testing regardless of geography or proximity to central laboratories. Moreover, advanced molecular laboratory infrastructure available in central reference laboratories can be utilized for blood donation screening – an important component of HCV elimination efforts.^8^ High prevalence of anti-HCV antibodies among blood donors is well documented in Pakistan.^42,43^ Available infrastructure could also be deployed in other disease elimination programs, including HIV, HBV, *Mycobacterium tuberculosis*, human papillomavirus, and SARS-CoV-2.

A limited number of economic evaluations of HCV elimination diagnostic strategies have been previously conducted in Pakistan.^13,44^ Chhatwal et al. investigated the cost of HCV elimination in Pakistan using different combinations of tests for screening and confirmation of viremia.^13^ A testing strategy involving the use of a POC screening test followed by the GeneXpert test for detection of viremia and assessment of treatment response yielded the lowest annual cost. This finding is different to this study and can be attributed to differences in the study design and application of different costing parameters. Other HCV-related economic evaluation studies with relevance to Pakistan did not assess the cost-effectiveness of a comprehensive array of HCV diagnostic testing approaches.^15,44-46^

The WHO has set an ambitious goal that would require the diagnosis of 90% of all patients with HCV and treatment initiation for 80% of eligible patients with HCV by the year 2030.^8^ Other targets include a 90% reduction in incident cases and a 65% reduction in HCV-associated mortality. A recent Pakistani study indicated that HCV elimination may indeed be feasible,^13^ and the Pakistani government has communicated a bold vision, committing to eliminate HCV infection by 2030. Initiatives such as the Prime Minister’s Hepatitis C Control Program and the establishment of the National Blood Transfusion Authority in Pakistan are encouraging in that regard.^47^

Our study findings have potential health policy implications in Pakistan. Given that, in Pakistan, healthcare expenditure makes up only 0.9% of gross domestic product^48^ and budgets are therefore severely constrained, our analysis provides decision-makers with evidence on the optimal approach to scale up HCV screening. Our analysis suggests that a centralized reference laboratory testing approach supplemented with novel sample collection methods, such as the cobas PSC, would provide the best value for money in Pakistan. In line with WHO and European Association for the Study of the Liver HCV testing and treatment guidelines,^49,50^ there is a possibility to innovate further and allow ‘real-reflex’ testing with the PSC. Two spots could be sampled on the PSC to enable laboratory-based anti-HCV testing and subsequent confirmatory testing (e.g., on the cobas HCV test for use with cobas 6800/8800 systems) without the need for additional sampling.

In developing HCV testing and scale-up plans, a comprehensive accounting of all costs along the entire continuum of a patient testing journey is warranted to objectively inform resource allocation decisions. Focusing solely on the price of the consumables may significantly underestimate the full cost burden associated with testing. High-throughput testing that is synergized with highly accurate assays can leverage the economies of scale to support expanded access to HCV testing at lower costs.^51^ Fully automated high-throughput solutions can play a key role in rapidly scaling up testing and can accelerate streamlined linkage to care and treatment. Egypt is poised to become the first country to eliminate HCV. This success is partly attributed to availability of high-throughput molecular diagnostic solutions.^52^ Testing platforms have now been repurposed for donor blood screening and COVID-19 pandemic testing.

Our study is subject to several limitations. First, there is a lack of head-to-head test performance studies that directly compare POC confirmatory testing with reference laboratory testing using the PSC. The model test performance inputs of POC NAT testing and central NAT testing were based on a synthesis of results of evaluations of the performance under different field conditions. An analysis including the DBS approach (a potential alternative to the cobas PSC) was beyond the scope of this study. However, deterioration of DBS viral recovery has been widely discussed in HIV and HCV studies,^23,25,53,54^ whereas PSC has been demonstrated to exhibit high stability up to 4 weeks.^21^ Second, we did not account for potential societal costs, including patient transport costs to the GeneXpert testing locations and related productivity losses associated with time off from work. We also did not consider the costs associated with returning results to patients following laboratory-based confirmatory testing. We assumed that these costs would be negligible. Third, we did not capture the long-term downstream impact of testing, including potential reduction in onward disease transmission. We postulate that expanded access to testing followed by appropriate treatment would curtail disease transmission among patients achieving sustained virologic response.

## CONCLUSIONS

Given the high prevalence of HCV infection in Pakistan and the humanistic burden of illness associated with chronic HCV, a focus on expanding screening programs and linkage to care is critical toward meeting WHO 2030 elimination targets. The base case results from this study suggest that a reference laboratory-based approach would provide the best value for money when scaling up HCV screening in Pakistan. High-throughput centralized testing can rapidly expand access. Additionally, this analysis underscores the value of novel sample collection technologies, such as the cobas PSC, which may help overcome challenges associated with rural population testing, LTFU, and specimen transport. It also demonstrates that an HCV screening approach that assumes the use of the cobas PSC to collect blood samples for confirmatory testing is highly likely to be cost effective compared to a near-patient molecular POC approach. The cobas PSC has the potential to significantly increase access to HCV testing in ‘hard-to-reach’ rural areas in Pakistan and may play an essential role in helping other countries to scale up testing to meet WHO HCV elimination goals.

## Data Availability

All data produced in the present study are available upon reasonable request to the authors

## Abbreviations

Ab: antibody
CHEERS: Consolidated Health Economic Evaluation Reporting Standards
CEN: central laboratory testing
CI: confidence interval
DAA: direct-acting antiviral agent
DBS: dried blood spot
EC: electric consumption
FN,: false negative
FP: false positive
HCV: hepatitis C virus
HIV: human immunodeficiency virus
ICER: incremental cost-effectiveness ratio
kWa: kilowatt hour
LMIC: low- and middle-income country
LTFU: lost to follow-up
MOH: Ministry of Health
NAT: nucleic acid test
P: probability
POC: point of care
PSA: probabilistic sensitivity analysis
PSC: Plasma Separation Card
RDS: Roche Diagnostic Systems
RNA: ribonucleic acid
TN: true negative
TP: true positive
US: United States
VA: volt-ampere
WHO: World Health Organization

## Acknowledgements

We thank Dr Huma Qureshi and Dr Hassan Mahmood for providing local Pakistan data and expert insights that guided the design of this study. This study was funded by Roche Molecular Systems, Inc. Editorial support with the preparation of the manuscript was provided by Tina Patrick at Elements Communications Ltd., Westerham, UK and was funded by Roche Molecular Systems, Inc.

## Competing interests

JBB and SC served as paid consultants to Roche Molecular Systems, Inc. JKK is an employee of Roche Diagnostics Solutions. MBM is an employee of Roche Molecular Systems, Inc. MMC was an employee of Roche Molecular Systems, Inc. at the time this study was conducted and remains a stockholder. LPG served as a paid consultant to Roche Molecular Systems, Inc. and received grants or has contracts with Gilead Sciences. SSH has no conflicts of interest to declare.

COBAS is a trademark of Roche. All other product names and trademarks are the property of their respective owners.

## Authors’ contributions

JBB, SC, and LPG led the study design. JBB, JKK, MBM, and MMC prepared the draft manuscript. All authors reviewed the analysis, contributed to the interpretation of data, and provided substantial modifications to the final manuscript.

